# COVID-19 Case Mortality Rates Continue to Decline in Florida

**DOI:** 10.1101/2020.08.03.20167338

**Authors:** Jeffrey E. Harris

**Affiliations:** Professor of Economics, Massachusetts Institute of Technology, Cambridge MA 02139

**Keywords:** SARS-CoV-2, coronavirus, case fatality rate, incidence rate, hospitalization, case severity, learning by doing, age groups, congestion

## Abstract

We studied COVID-19 case mortality in Florida among four successive cohorts of persons at least 50 years of age, each of whom we followed for 28 to 48 days from date of diagnosis. The cohorts were separated by date of diagnosis into four nonoverlapping intervals: March 29 - April 18; April 29 - May 19; May 21 - June 10; and June 14 - July 4, 2020. Case mortality rates declined consistently and significantly over the course of the four intervals: 57% among those aged 50-59 years; 62% among those aged 60-69 years; 52% among those aged 70-79 years; and 34% among those aged 80 or more years. These findings were consistent with progressive improvements in the medical care of COVID-19 patients. We further studied case mortality by hospitalization status. The case mortality rate among hospitalized patients aged 60-69 years fell significantly from the first to the third interval. During the fourth interval, an apparent rise in mortality among hospitalized patients in the same age group was mirrored by a significant decline in mortality among those not hospitalized. These findings were consistent with the out-of-hospital treatment of some patients who would have previously been hospitalized.

This study relies exclusively on publicly available, aggregate health data that contain no individual identifiers. The author has no competing interests and no funding sources to declare. This article represents to the sole opinion of its author and does not necessarily represent the opinions of the Massachusetts Institute of Technology, the National Bureau of Economic Research, Eisner Health, or any other organization.

## Introduction

Numerous investigators have studied the cross-sectional relationship between COVID-19 mortality and patient age, gender, comorbid medical conditions, and various biomarkers (Palaiodimos et al. 2020, Tian et al. 2020, Wang et al. 2020). Few authors, however, have systematically studied longitudinal trends in COVID-19 case mortality. One study found improvements in hospital-based mortality in a relatively small cohort (Ciceri et al. 2020). Another group attributed the recent decline in hospital-based mortality in the United Kingdom to various factors, including changing patient age distribution and case severity, as well as improved clinical management (Mahon, Oke, and Heneghan 2020).

Studies of trends in COVID-19 case mortality rates in the general population, including non-hospitalized patients, appear to be even scarcer. Part of the difficulty is the lead time between onset of symptoms and ultimate death, estimated to be on the order of 16 days (Harris 2020a, Muzimoto and Chowell 2020). Without adequate follow up of initially diagnosed cases, data on death outcomes will be subject to right truncation. We could falsely conclude that death rates are falling when we simply haven’t waited long enough to see who has died (Harris 2020b).

Here, we analyze a large database of individual COVID-19 cases, made publicly available by the Florida Department of Public Health, to analyze recent trends in case mortality rates among persons aged 50 years or more.

## Methods

### Data

We downloaded a file named *Florida_COVID19_Case_Line_Data*.*csv* from the Florida Department of Public Health website (Florida Department of Public Health 2020) on four distinct dates: (1) May 16, 2020; (2) June 16, 2020; (3) July 8, 2020; and (4) August 1, 2020. The file downloaded on each date included a separate record for each case of diagnosed COVID-19 thus far reported. Each record included (if known) the individual’s age, vital status as of the download date, and whether the individual had been hospitalized at any time up to the download date. The counts of individuals aged 50 years or more were, respectively: (1) 23,152 (51.7%) out of 44,811 cases reported through May 16, 2020; (2) 35,404 (44.2%) of 80,109 cases reported through June 16, 2020; (3) 76,298 (34.1%) out of 223,783 cases reported through July 8, 2020; and (4) 167,231 (34.8%) of 480,028 cases reported through August 1, 2020.

For each downloaded file, we defined a distinct, nonoverlapping diagnostic interval during which cases were eligible for follow up: (1) March 29 – April 18 for cases reported through May 16, 2020; (2) April 29 – May 19 for cases reported on June 16, 2020; (3) May 21 – June 10 for cases reported through July 8, 2020; and (4) June 14 – July 4 for cases reported through August 1, 2020. We selected these cutoff dates so that each individual diagnosed during an interval would be effectively subject to a mortality follow up of 28 to 48 days. Consider, for example, those COVID-19 cases diagnosed during the first interval, whose vital status was known as of May 16, 2020. An individual diagnosed on the first day of the interval (March 29) would have a vital status follow up of 48 days, while an individual diagnosed on the last day of the interval (April 18) would have a vital status follow up of 28 days. We thus had four mutually exclusive cohorts of COVID-19 cases, which were approximately equally separated over time. Each cohort consisted entirely of individuals aged 50 years or more, and each cohort had the same range of mortality follow-up.

### Statistical Analysis

We classified cases by diagnostic interval, 10-year age group (50–59 years, 60–69 years, 70–79 years, or 80 or more years), hospitalization status (hospitalized, not hospitalized, or unknown) and vital status (alive or dead). The Florida Department of Public Health did not report any COVID-19 case as having unknown vital status.

We ran two types of analyses. In the first aggregated analysis, we calculated trends in case mortality by 10-year age group over time, that is, over the four diagnostic intervals. For each age group and each diagnostic interval, we calculated the case mortality rate and associated 95 percent confidence interval based on the exact binomial distribution. We ran Poisson regressions to assess the rate of change per time interval, measuring each interval as an integer from 1 to 4.

In the second disaggregated analysis, we calculated trends in case mortality by 10-year age group and hospitalization status over time. Using graphical methods, we focused sharply on trends in case mortality of hospitalized and non-hospitalized patients aged 60–69 years.

## Results

### Aggregate Analysis by Age Group

Figure 1 displays the principal results of our aggregate analysis by ten-year age group. Each color corresponds to a different age group: 50–59 years (lime), 60–69 years (pink), 70–79 years (mango), and 80 years or more (cyan). The data points represent case mortality rates, as gauged on a logarithmic scale, shown at the left. The vertical bars surrounding each data point represent the 95 percent confidence intervals. The mortality estimates are arrayed in vertical columns, each column corresponding to one of the four diagnostic intervals.

**Figure 1.**
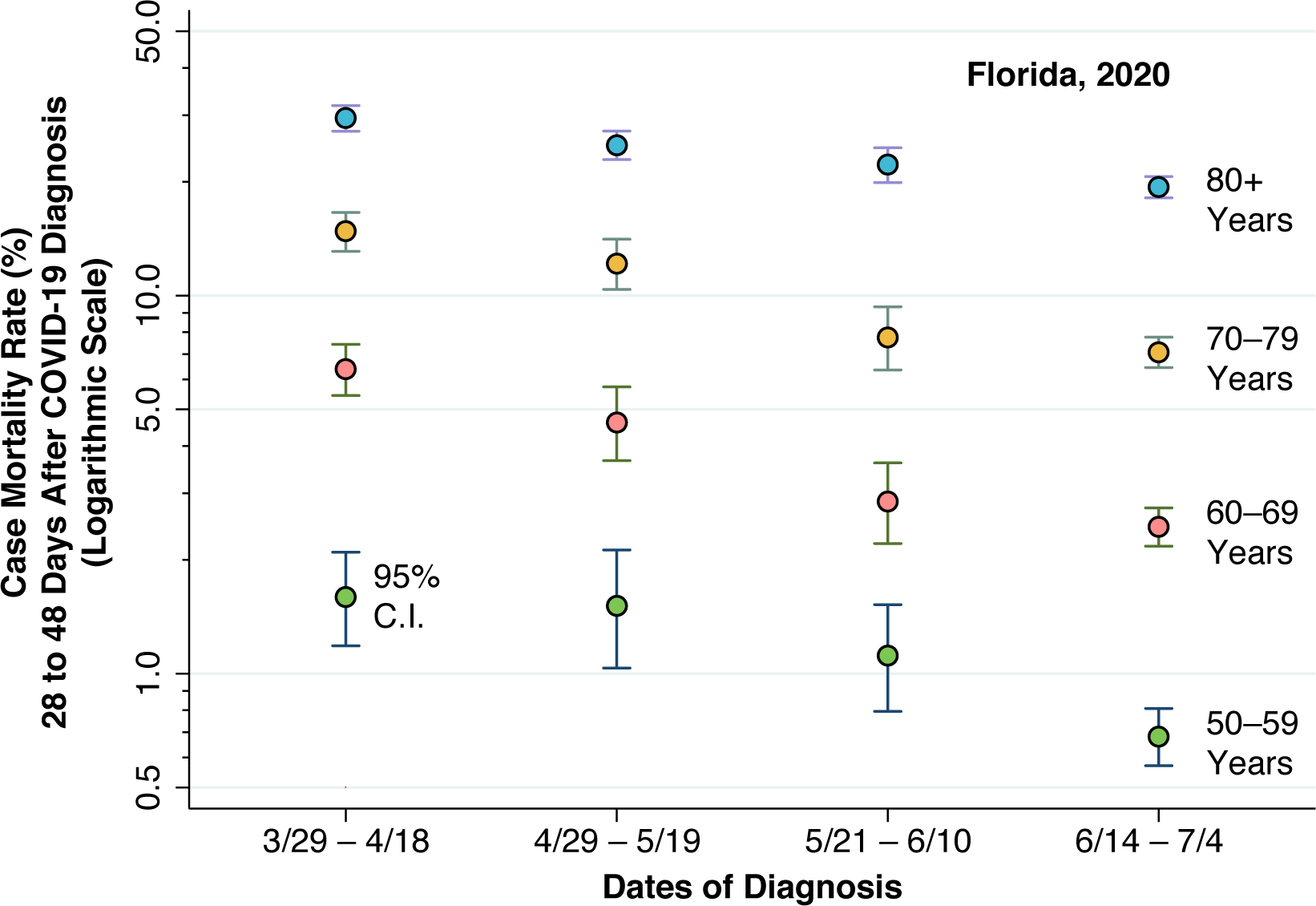
Case Mortality Rate 28 to 48 Days After COVID-19 Diagnosis by Age Group During Four Nonoverlapping Diagnostic Intervals, Florida, 2020

Figure 1 shows that case mortality has been steadily declining in Florida since the end of March 2020 in all four age groups. Trend regression analyses showed significant rates of decline: 30.1 percent per diagnostic interval (95% confidence interval (C.I.) 20.1–40.1, *P* < 0.001) among persons 50–59 years; 32.1 percent per diagnostic interval (95% C.I. 25.8–38.5, *P* < 0.001) among persons 60–69 years; 25.1 percent per diagnostic interval (95% C.I. 20.1–30.2, *P* < 0.001) among persons 70–70 years; and 13.8 percent per diagnostic interval (95% C.I. 10.1–17.5, *P* < 0.001) among persons 80 years or more. The overall declines in case mortality in the three months from the beginning of the first interval to the end of the fourth interval were: 57% among those aged 50-59 years; 62% among those aged 60-69 years; 52% among those aged 70-79 years; and 34% among those aged 80 or more years

### Disaggregated Analysis by Age Group and Hospitalization Status

Appendix Table 1 shows the detailed results of our disaggregated analysis by age group and hospitalization status. Figure 2 focuses specifically on the age group 60–69 years. As in Figure 1, each data point and surrounding vertical bars represent an estimated case mortality rate with 95 percent confidence interval. (We have dropped some half-bars for visual clarity.) Similarly, as in Figure 1, the case mortality rates are gauged in a logarithmic scale, as shown on the left-hand axis. Also, as in Figure 1, the vertical columns correspond to the successive diagnostic intervals over time.

**Figure 2.**
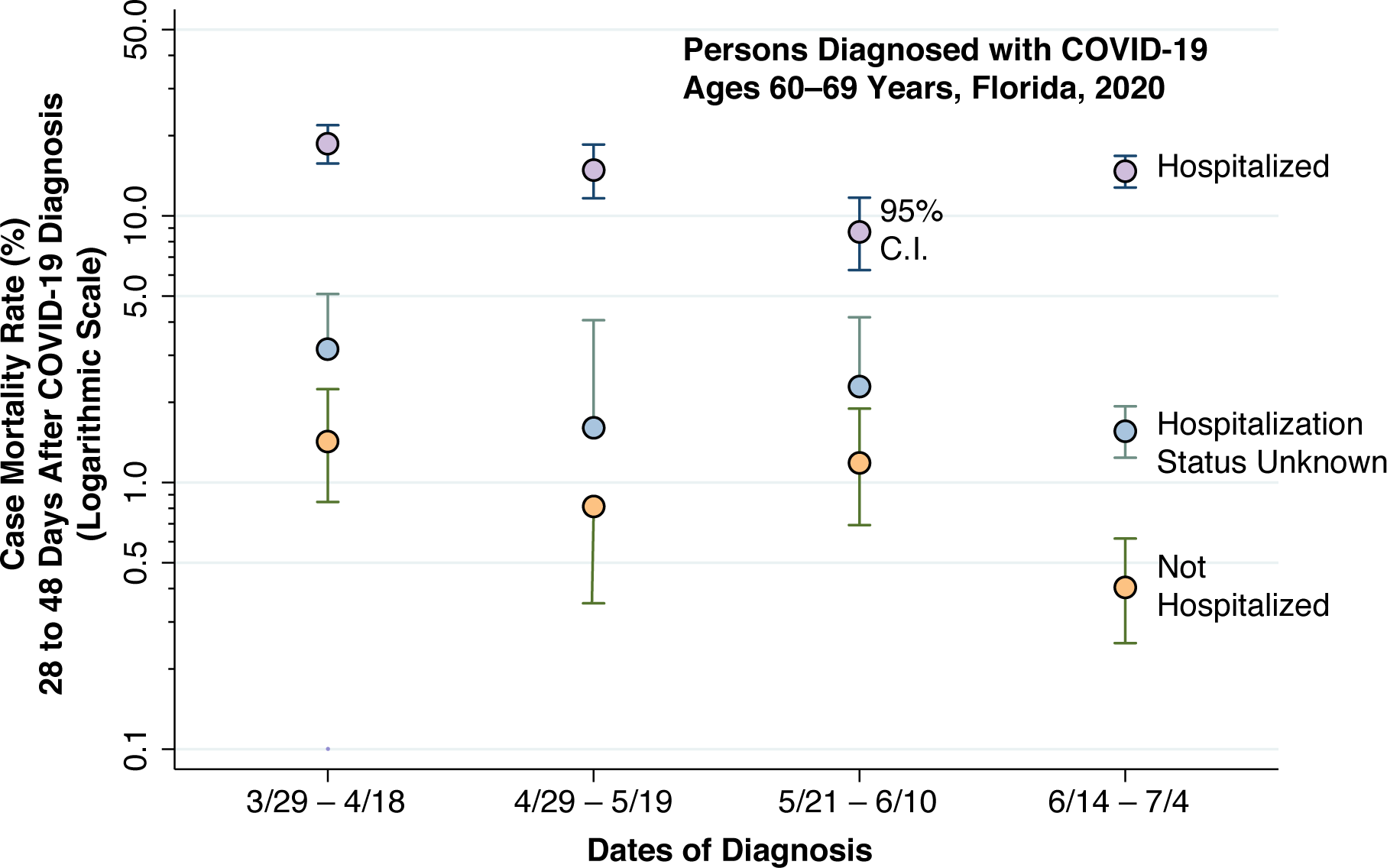
Case Mortality Rate 28 to 48 Days After COVID-19 Diagnosis by Hospitalization Status Among Persons Aged 60–69 Years During Four Nonoverlapping Diagnostic Intervals, Florida, 2020

In contrast to Figure 2, the different colors correspond to varying hospitalization status: hospitalized (lilac), unknown status (sky blue), and not hospitalized (peach). Among hospitalized patients, there is an initial decline during the first three diagnostic intervals, followed by an apparent rebound in case mortality. By contrast, among both non-hospitalized individuals and those with unknown hospitalization status, the trends during the second, third and fourth diagnostic intervals appear to follow a mirror image. More specifically, as case mortality rebounded in hospitalized patients during the last diagnostic interval, case mortality among non-hospitalized individuals and those with unknown status declined. While there is more variability in the estimated case mortality rates, the same underlying pattern can be seen in the other age groups, as evidenced in Appendix Table 1.

## Discussion

This longitudinal study of individuals diagnosed with COVID-19 in the state of Florida shows a substantial, continuing decline in case mortality rates among individuals aged 50 years or more. The decline was observed in every ten-year age group, from those aged 50–59 years to those aged 80 years or more. Focusing on those aged 60–69 years old, we observed a decline in case mortality among hospitalized patients during the first three diagnostic intervals (March 29– April 18, April 29–May 19, and May 21–June 10), but a rebound in case mortality in the final diagnostic interval (June 14–July 4).

A lack of detailed, individual-level clinical data makes it difficult to distinguish with high confidence between three potential, but not mutually exclusive explanations for the observed findings. First, the marked decline in case mortality may have been due to increasing, systematic under-ascertainment of death. Second, there may have been a major shift in case mix, with less severe cases increasingly dominant. Third, clinical care of the sickest COVID-19 patients may have markedly improved. Based upon the available evidence, however, the latter explanation is far and away the most plausible.

We have found no evidence that systematic underreporting of deaths could quantitatively explain progressive declines in mortality of the magnitude seen here. Possible changes in case mix would be more a concern if we had not excluded younger persons under 50 or had not controlled for age. Analyses of Florida testing data go against the hypothesis that a massive increase in testing stimulated the detection of milder cases that would have otherwise gone undetected (Harris 2020b, c).

The significant decline in case mortality – a reduction of at least 34 percent over a three-month period in every ten-year age group over 50 years of age – is in all likelihood the result of genuine improvements in clinical care. Quite apart from new clinical approaches with concrete evidentiary support – including the more judicious use of high-flow oxygenation rather than mechanical ventilation, the turning of patients onto a prone or semi-prone position, the administration of prophylactic anticoagulants, high-dose corticosteroids and selected antiviral agents – advances in care are often less concretely embodied in what economists have described as learning by doing (Arrow 1962, Broulliette 2020). That includes learning what clinical practices to avoid. Improvements in the care of residents of assisted living facilities, which comprise a nontrivial fraction of Florida’s elderly population, could also have contributed (Grabowski and Mor 2020).

The finding that case fatality rates declined among hospitalized patients in their 60s, at least during the first three diagnostic intervals (Figure 2), is more consistent with genuine improvements in care than the hypothesis of progressively milder case severity. The observation that case mortality subsequently increased among hospitalized 60-year-olds raises the possibility of congestion effects (Yu et al. 2020). However, the finding of a coincident decline in case mortality among non-hospitalized 60-year-olds points instead to enhanced triaging of less seriously ill patients to out-of-hospital care. Not only has inpatient care improved, but healthcare providers have also come to understand better when patients do and do not need hospitalization.

The present finding of a continued decline in COVID-19 case mortality, however, does not necessarily translate into a corresponding decline in population mortality rates. Population mortality is the product of two variables: disease incidence and case fatality. Unfortunately, even among the elderly, the incidence of new COVID-19 cases has recently increased in Florida at a rate faster than the decline in case mortality reported here (Harris 2020b, COVID Tracking Project 2020).

## Data Availability

The underlying data links are provided in the article. The author will make all intermediate programs and output available.

---

**Appendix Table 1.**

| Diagnosis Interval | Hospitalization Status | Age Group | Number of Deaths | Total Cases | Mortality Rate (%) | Lower Bound (%) | Upper Bound (%) |
| --- | --- | --- | --- | --- | --- | --- | --- |
| 3/29–4/18 | No | 50-59 | 9 | 2,016 | 0.4 | 0.2 | 0.8 |
| 3/29–4/18 | No | 60-69 | 18 | 1,265 | 1.4 | 0.8 | 2.2 |
| 3/29–4/18 | No | 70-79 | 17 | 637 | 2.7 | 1.6 | 4.2 |
| 3/29–4/18 | No | 80+ | 68 | 476 | 14.3 | 11.3 | 17.8 |
| 3/29–4/18 | Yes | 50-59 | 37 | 494 | 7.5 | 5.3 | 10.2 |
| 3/29–4/18 | Yes | 60-69 | 120 | 643 | 18.7 | 15.7 | 21.9 |
| 3/29–4/18 | Yes | 70-79 | 206 | 699 | 29.5 | 26.1 | 33 |
| 3/29–4/18 | Yes | 80+ | 348 | 816 | 42.6 | 39.2 | 46.1 |
| 3/29–4/18 | Unknown | 50-59 | 4 | 628 | 0.6 | 0.2 | 1.6 |
| 3/29–4/18 | Unknown | 60-69 | 16 | 505 | 3.2 | 1.8 | 5.1 |
| 3/29–4/18 | Unknown | 70-79 | 20 | 308 | 6.5 | 4 | 9.9 |
| 3/29–4/18 | Unknown | 80+ | 40 | 255 | 15.7 | 11.4 | 20.7 |
| 4/29–5/19 | No | 50-59 | 5 | 1,462 | 0.3 | 0.1 | 0.8 |
| 4/29–5/19 | No | 60-69 | 8 | 982 | 0.8 | 0.4 | 1.6 |
| 4/29–5/19 | No | 70-79 | 31 | 602 | 5.1 | 3.5 | 7.2 |
| 4/29–5/19 | No | 80+ | 87 | 606 | 14.4 | 11.7 | 17.4 |
| 4/29–5/19 | Yes | 50-59 | 26 | 318 | 8.2 | 5.4 | 11.8 |
| 4/29–5/19 | Yes | 60-69 | 65 | 438 | 14.8 | 11.6 | 18.5 |
| 4/29–5/19 | Yes | 70-79 | 112 | 463 | 24.2 | 20.4 | 28.4 |
| 4/29–5/19 | Yes | 80+ | 261 | 703 | 37.1 | 33.5 | 40.8 |
| 4/29–5/19 | Unknown | 50-59 | 1 | 340 | 0.3 | 0 | 1.6 |
| 4/29–5/19 | Unknown | 60-69 | 4 | 249 | 1.6 | 0.4 | 4.1 |
| 4/29–5/19 | Unknown | 70-79 | 8 | 178 | 4.5 | 2 | 8.7 |
| 4/29–5/19 | Unknown | 80+ | 43 | 254 | 16.9 | 12.5 | 22.1 |
| 5/21–6/10 | No | 50-59 | 4 | 2,431 | 0.2 | 0 | 0.4 |
| 5/21–6/10 | No | 60-69 | 17 | 1,433 | 1.2 | 0.7 | 1.9 |
| 5/21–6/10 | No | 70-79 | 16 | 672 | 2.4 | 1.4 | 3.8 |
| 5/21–6/10 | No | 80+ | 35 | 402 | 8.7 | 6.1 | 11.9 |
| 5/21–6/10 | Yes | 50-59 | 23 | 448 | 5.1 | 3.3 | 7.6 |
| 5/21–6/10 | Yes | 60-69 | 39 | 448 | 8.7 | 6.3 | 11.7 |
| 5/21–6/10 | Yes | 70-79 | 70 | 380 | 18.4 | 14.7 | 22.7 |
| 5/21–6/10 | Yes | 80+ | 205 | 544 | 37.7 | 33.6 | 41.9 |
| 5/21–6/10 | Unknown | 50-59 | 12 | 617 | 1.9 | 1 | 3.4 |
| 5/21–6/10 | Unknown | 60-69 | 10 | 437 | 2.3 | 1.1 | 4.2 |
| 5/21–6/10 | Unknown | 70-79 | 15 | 251 | 6 | 3.4 | 9.7 |
| 5/21–6/10 | Unknown | 80+ | 36 | 298 | 12.1 | 8.6 | 16.3 |
| 6/14–7/4 | No | 50-59 | 14 | 9,137 | 0.2 | 0.1 | 0.3 |
| 6/14–7/4 | No | 60-69 | 21 | 5,204 | 0.4 | 0.2 | 0.6 |
| 6/14–7/4 | No | 70-79 | 46 | 2,320 | 2 | 1.5 | 2.6 |
| 6/14–7/4 | No | 80+ | 105 | 1,100 | 9.5 | 7.9 | 11.4 |
| 6/14–7/4 | Yes | 50-59 | 82 | 1,160 | 7.1 | 5.7 | 8.7 |
| 6/14–7/4 | Yes | 60-69 | 183 | 1,245 | 14.7 | 12.8 | 16.8 |
| 6/14–7/4 | Yes | 70-79 | 266 | 1,134 | 23.5 | 21 | 26 |
| 6/14–7/4 | Yes | 80+ | 439 | 1,130 | 38.8 | 36 | 41.8 |
| 6/14–7/4 | Unknown | 50-59 | 35 | 8,908 | 0.4 | 0.3 | 0.5 |
| 6/14–7/4 | Unknown | 60-69 | 81 | 5,201 | 1.6 | 1.2 | 1.9 |
| 6/14–7/4 | Unknown | 70-79 | 117 | 2,600 | 4.5 | 3.7 | 5.4 |
| 6/14–7/4 | Unknown | 80+ | 204 | 1,633 | 12.5 | 10.9 | 14.2 |

